# Predictive validity of a DNA methylation-based screening panel for postpartum depression

**DOI:** 10.1101/2020.03.05.20027847

**Authors:** Dana M. Lapato, Roxann Roberson-Nay, Patricia A. Kinser, Timothy P. York

## Abstract

Prenatal maternal depression increases the risk of negative maternal-infant health outcomes but often goes unrecognized. As a result, biomarker screening tests capable of identifying women at risk for depression are highly desirable. This study tested how demographic and clinical factors affect the predictive validity of a DNA methylation-based screening test for postpartum major depression (MD) using data from a longitudinal study of birth outcomes. Lifetime history of MD and current levels of postpartum depressive symptoms were assessed using an extended self-report version of the Composite International Diagnostic Interview Short Form and the Edinburgh Postnatal Depression Scale (EPDS), respectively. Predictive validity of the test was estimated in the PREG cohort using the area under the receiver operator characteristic curve (AUC), and sensitivity analyses were performed to assess the impact of self-reported race, age, and pre-pregnancy history of MD. Data for N=103 pregnant participants (African-American=49; European-American=54) were available. The prediction model identified women who would develop high levels of postpartum depressive symptoms better within the subset of women with previous histories of MD (AUC = 0.94, 95% CI 0.79-1.00) compared to the full pregnant cohort (AUC = 0.62, 95% CI 0.46-0.79). This observation prompted secondary analyses to test the model specificity for postpartum depression. The model predicted lifetime history of MD moderately well in never-pregnant, mixed-sex cohort of adolescents (N=150; ages 15-20; AUC = 0.75, 95% CI 0.57-0.92) and performed slightly better in males versus females. Additional sensitivity analyses are needed to determine the extent of the model’s specificity for MD subtypes and if demographic or clinical factors influence the predictive validity of this model.

## 1 Introduction

Maternal depression is one of the most prevalent complications associated with childbirth. ^1^ Untreated maternal depression during pregnancy or shortly following birth increases the risk of serious adverse pregnancy, maternal, and infant-child outcomes, even if the depressive symptoms do not reach established diagnostic thresholds. ^1,2,3,4^ Treatment for maternal depression during pregnancy can significantly reduce the odds of negative health consequences. ^4,5^ As a result, identifying pregnant women at elevated risk for or currently experiencing maternal depression is a critical focal point for developing perinatal health interventions. Efforts to raise awareness of perinatal-onset MD have included the addition of a specifier to the fifth edition of the Diagnostic and Statistical Manual (DSM-5) and updated perinatal depression screening guidelines from the American College of Obstetricians and Gynecologists (ACOG). Despite these changes, recognizing MD episodes and depressive symptoms during pregnancy and the early postpartum remains challenging. Patients and clinicians alike may struggle to determine if the constellation of symptoms reflects classic somatic indicators of MD (e.g., changes in weight/appetite, difficulty sleeping, and fatigue) or benign pregnancy-related physiological and/or hormonal changes. ^6^

Given the challenges of assessing maternal perinatal depressive psychopathology, biomarker-based screening tests capable of indexing risk for perinatal-onset MD could prove invaluable for reducing the number of unrecognized cases of maternal depression. DNA methylation (DNAm) has become a prominent biomarker candidate, in part because it is easily measured in peripheral blood and its patterning can be influenced by both genetic and environmental factors. ^7,8,9,10^ DNAm is a chemical modification to DNA that can influence gene expression and genomic stability without altering the DNA sequence. In 2014, two DNAm sites near *TTC9B* and *HP1BP3* were associated with postpartum depression (PPD) and acheived high levels of clinical predictive accuracy. ^11^ Follow-up analyses of related biomarkers have been performed in other cohorts, ^12^ including assessments of gene expression and postpartum DNAm remodeling trajectories of the loci near *TTC9B* and *HP1BP3*. Due to the potential importance of the original finding, replication studies in independent cohorts and additional sensitivity analyses should be performed to assess the generalizability of the prediction algorithm. So far, the tested cohorts have been composed predominantly of participants who self-identified as Caucasian, were in their early 30’s, and had a significant histories of either MD or bipolar disorder (see Table 2). As a result, the impact of genetic ancestry, maternal age, and maternal history of psychopathology on the predictive validity of the DNAm biomarkers is unclear.

**Table 1:**
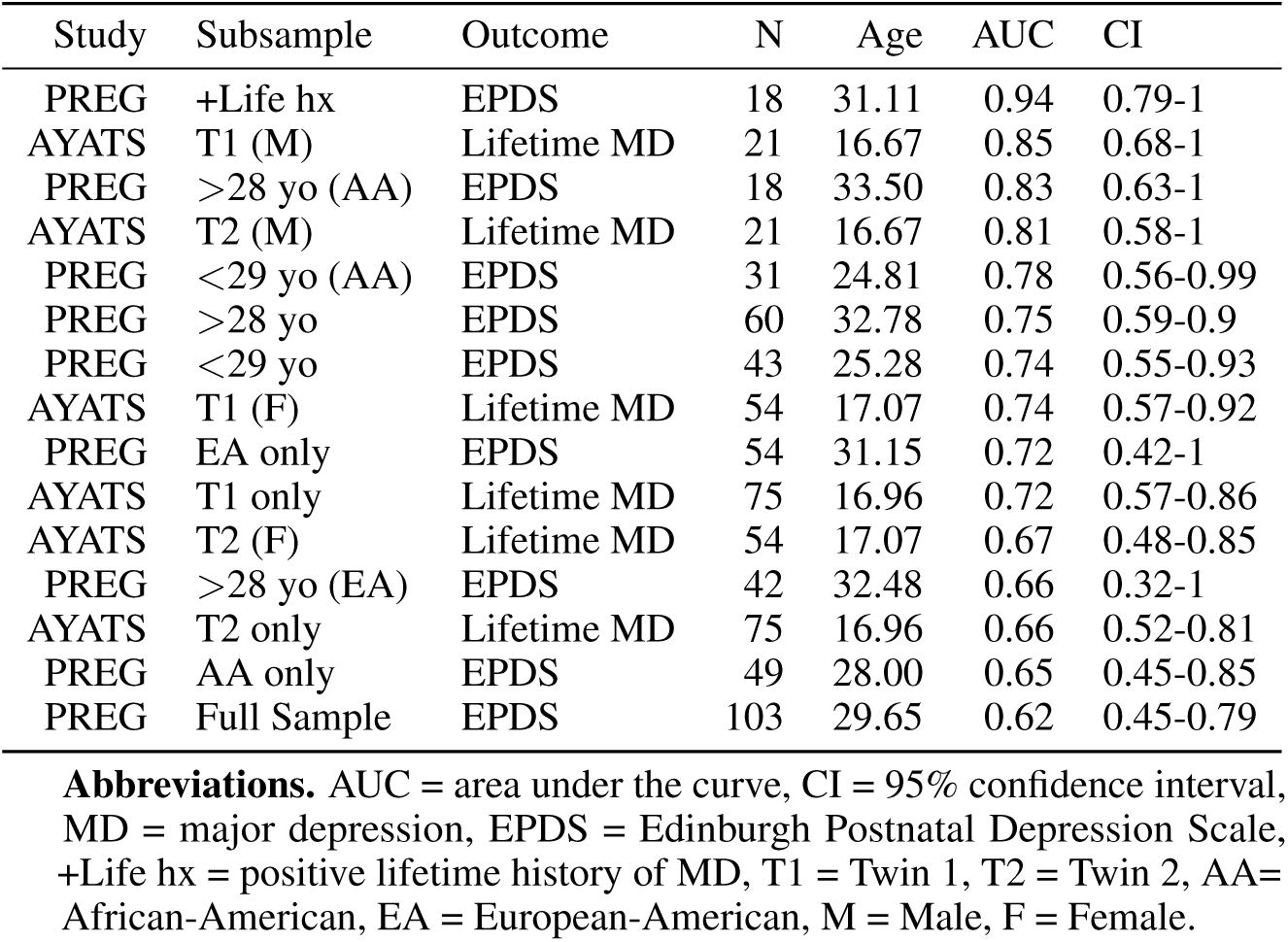
Model Results

| Study | Subsample | Outcome | N | Age | AUC | CI |
| --- | --- | --- | --- | --- | --- | --- |
| PREG | +Life hx | EPDS | 18 | 31.11 | 0.94 | 0.79-1 |
| AYATS | T1 (M) | Lifetime MD | 21 | 16.67 | 0.85 | 0.68-1 |
| PREG | >28 yo (AA) | EPDS | 18 | 33.50 | 0.83 | 0.63-1 |
| AYATS | T2 (M) | Lifetime MD | 21 | 16.67 | 0.81 | 0.58-1 |
| PREG | <29 yo (AA) | EPDS | 31 | 24.81 | 0.78 | 0.56-0.99 |
| PREG | >28 yo | EPDS | 60 | 32.78 | 0.75 | 0.59-0.9 |
| PREG | <29 yo | EPDS | 43 | 25.28 | 0.74 | 0.55-0.93 |
| AYATS | T1 (F) | Lifetime MD | 54 | 17.07 | 0.74 | 0.57-0.92 |
| PREG | EA only | EPDS | 54 | 31.15 | 0.72 | 0.42-1 |
| AYATS | T1 only | Lifetime MD | 75 | 16.96 | 0.72 | 0.57-0.86 |
| AYATS | T2 (F) | Lifetime MD | 54 | 17.07 | 0.67 | 0.48-0.85 |
| PREG | >28 yo (EA) | EPDS | 42 | 32.48 | 0.66 | 0.32-1 |
| AYATS | T2 only | Lifetime MD | 75 | 16.96 | 0.66 | 0.52-0.81 |
| PREG | AA only | EPDS | 49 | 28.00 | 0.65 | 0.45-0.85 |
| PREG | Full Sample | EPDS | 103 | 29.65 | 0.62 | 0.45-0.79 |

**Abbreviations.** AUC = area under the curve, CI = 95% confidence interval, MD = major depression, EPDS = Edinburgh Postnatal Depression Scale, +Life hx = positive lifetime history of MD, T1 = Twin 1, T2 = Twin 2, AA= African-American, EA = European-American, M = Male, F = Female.

**Table 2:** Comparison of Current Analysis with Previous Studies of the Postpartum Depression Predictive Algorithm

| Cohort | N | Age, M(sd) | Race | MD/BP (%) | DNAm timeframe | Postpartum MD/Dsx | Instrument |
| --- | --- | --- | --- | --- | --- | --- | --- |
| PREG PPD | 103 | 29.6 (4.5) | 52.4% Caucasian<br>47.6% African American | 17.5 / * | Early pregnancy | ≤ 7 months | EPDS |
| AYATS | 150 | 17.5 (1.28) | 100% Caucasian | 26 / 0 | Nulligravida | NA | CIDI-SF<sup>1</sup> |
| JHU PPD Cohort | 93<sup>2</sup> | 30.6 (6.3) | 70% Caucasian | ~66 / ~33 | Prenatal | 1 month | Psychiatric interview |
| GenRED | 84 | 39.7 (.09) | 92% Caucasian<br>1.2% African American<br>1.2% Asian<br>3.6% Unreported | 44.6 / 2.4 | Postpartum;<br>Time since delivery unclear | Unclear | Unclear |
| FRAMES | 240 | 32.7 (.02) | 100% Caucasian | * / * | 1-3 years postpartum | 6-8 months | HDRS |
| Emory PPD Cohort | 62 | 33 | ~85% Caucasian | 59.6 / 40.6 | Prenatal | Postpartum | HDRS |

\* Not assessed
<sup>1</sup> An extended self-report version of the CIDI-SF was used to assess lifetime history of MD.
Race is based on self-identified Census-based race categories. DNAm and Postpartum MD/Dsx columns show time of blood sample collection and psychiatric assessment, respectively, relative to birth.
<sup>2</sup> The Guintivano (2014) paper describes the cohort as including 93 recruited women and analyses that involved 51 women. To the best of our knowledge, the clinical and demographic proportions listed in the paper refer to the recruited cohort.
**Abbreviations.** MD = major depression; BP = bipolar disorder; DNAm = DNA methylation; Dsx = depressive symptoms; NA = Not applicable; PREG PPD = Pregnancy, Race, Environment, Genes Study Postpartum Extension; EPDS = Edinburgh Postnatal Depression Scale; AYATS=Adolescent and Young Adult Twin Study; CIDI-SF = Composite International Diagnostic Interview-Short Form; JHU PPD Cohort = Johns Hopkins Prospective Postpartum Cohort<sup>11</sup>, GenRED=Genetics of Recurrent Early Onset Depression; FRAMES = Franconian Maternal Health Evaluation Studies; Emory PPD Cohort=Prospective Gene Expression Postpartum Cohort from the Emory Women's Mental Health Program<sup>34</sup>

The current study builds on initial findings by assessing the predictive validity and generalizability of the DNAm biomarker model in an epidemiological cohort of pregnant women from the Pregnancy, Race, Environment, Genes (PREG) study. ^13^ During the analysis, the high accuracy estimates were replicated for PPD risk, but only in a subset of participants with a lifetime history of MD. This observation motivated a specificity test of the prediction model to determine if the biomarkers associated uniquely with pregnancy-related depression. This analysis was carried out in a mixed-sex, never-pregnant subset of adolescent participants from the Adolescent and Young Adult Twin Study (AYATS).

## 2 Methods

### 2.1 Preregistation

The replication analysis was preregistered on the Open Science Framework (OSF) ^14^ using the AsPredicted format. The preregistration document and R code used to analyze the data and generate figures are available on the Open Science Framework (https://osf.io/7dsf9).

### 2.2 Samples

#### 2.2.1 Pregnancy, Race, Environment, Genes (PREG) Study

The Pregnancy, Race, Environment, Genes (PREG) Study was a prospective longitudinal study that followed approximately 230 women over the course of pregnancy. ^13^ Participants completed extensive questionnaires about lifetime and current exposure to social and environmental determinants of health up to four times during pregnancy. Peripheral blood was collected at each visit and used for DNAm analysis. Additional funding secured in the second year of the PREG study permitted enrolling approximately 100 of the participants into a postpartum extension that included two additional time points. Study enrollment criteria included 1) maternal age between 18 and 40 years old, 2) no use of artificial reproductive technology, 3) absence of diabetes, and 4) both parents had to self-identify as either European-American or African-American. Exclusion criteria at birth included any congenital, placental, or amniotic abnormalities (e.g., chromosomal abnormalities, polyhydramnios), preeclampsia/ PIH (pregnancy induced hypertension)/ HELLP (hemolysis, elevated liver enzymes, low platelet count), Rh sensitization, cervical cerclage, medically necessitated preterm delivery, drug abuse, and participating in fewer than 3 study time point assessments (including birth).

#### 2.2.2 Adolescent and Young Adult Twin Study (AYATS)

The Adolescent and Young Adult Twin Study (AYATS) is a longitudinal study that enrolled a general population sample of monozygotic (MZ) and dizygotic twin pairs between the ages of 15-20 years old. ^15^ Comprehensive questionnaires about mood and lifetime history of psychopathology were collected along with peripheral blood. DNAm was assayed on a subset of MZ twin pairs (N=75 pairs, N=150 twins, 73% female) selected based on pair status for lifetime MD (i.e., concordant positive, concordant negative, and discordant). No participants selected for DNAm measurement were using or had used antidepressant medication or medication with psychotropic effects for at least one month prior to providing a blood sample. Demographic information for the full AYATS sample and the DNAm subset are available elsewhere. ^15,16^

### 2.3 Depression phenotypes

*Lifetime history of MD* was measured using a self-report version of the Composite International Diagnostic Interview-Short Form (CIDI-SF). ^17^ In both the PREG study and AYATS, MD was considered present if at least four Criterion A symptoms, which had to include sad mood and/or anhedonia, were endorsed as present every day or nearly everyday, for the entire day or most of the day for two weeks. Additionally, participants had to endorse that the presence of the depression symptoms caused considerable distress or functional impairment. The use of a slightly lower threshold for defining clinical MD has been implemented previously and was justified given the that 1) participants must rely on retrospective memory and 2) that participants with four symptoms are phenotypically closer to a case (defined in the DSM-5 as the presence of at least five Criterion A symptoms in addition to interference ^18^) than a control. ^16,19,20^ Lifetime history of MD was assessed in both cohorts during the initial study visit. Use of antidepressant medication was not assessed in the PREG study.

*Current level of perinatal depressive symptoms* was assessed in PREG at postpartum study visits using the Edinburgh Postnatal Depression Scale (EPDS) between 1 and 3 months postpartum. ^21^ The EPDS is the most frequently used validated clinical tool to assess perinatal depressive symptoms. ^22,23^ Participants with EPDS scores of ≥13 were considered likely postpartum depression (PPD) cases. ^21,24^ Perinatal depressive symptoms were not assessed in AYATS because history of pregnancy was an exclusion criterion. ^15^

### 2.4 DNAm measurement and processing

Genome-wide DNAm was measured in both studies using the Illumina Infinium HumanMethylation450 microarray. Raw files were processed independently for each cohort in the R environment using Bioconductor packages. ^25,26,27,28^ Poor quality specimens were identified and removed before quantile normalization. Blood cell proportions were estimated using the Houseman method ^29^ consistent with the original study. ^11^ M-values were used for all analyses, and sensitivity analyses were performed with beta values to ensure that that data transformation did not affect the results. For all PREG participants, the DNAm sample from the first prenatal study visit was used since a previous study has shown the putative relationship between DNAm and postpartum depressive psychopathology is stronger in the first trimester compared to the third trimester. ^12^ ComBat was used to adjust for slide effects. ^30^

### 2.5 Statistical analysis

All analyses were performed in the R statistical environment using the pROC package. ^25,31^ The predictive formula from Guintivano (2014) is defined in Osborne (2016) as:

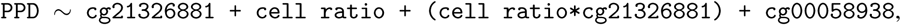

where PPD is a dichotomous outcome defined as the presence of postpartum MDP, cell ratio is the ratio of the estimated proportion of monocytes compared to the estimated proportions of CD4T cells, CD8T cells, B cells, and granulocytes (natural killer cells not listed as part of the calculation ^12^), and cg21326881 and cg00058938 representing the beta values of DNAm probe sites near the genes *HP1BP3* and *TTC9B*, respectively. The coefficients for the model were not reported in either paper.

Area under the receiver operator characteristic curves (AUC, ROC) was calculated for the full PREG sample to assess the model’s accuracy, and subsets of the PREG sample were modeled separately to test if the predictive validity of the model was influenced by demographic (e.g., age, self-identified Census-based race category) and clinical (e.g., lifetime history of depressive psychopathology) factors. Confidence intervals were calculated via bootstapping (k=2000).

Secondary analyses designed to test model specificity for pregnancy-related MD were structured so that lifetime history of MD (instead of perinatal depressive symptom load) was the predictor. The data used for this analysis came from AYATS. ^15^ To avoid bias from multiple genetically identical individuals, the AYATS sample was divided by twin order so that each group only had genetically distinct participants. The groups were further subdivided by sex to determine if the model could work in males.

## 3 Results

### 3.1 Study demographics

The PREG study had N=103 participants (African-American = 49; European-American = 54, mean age = 29.6 years (sd = 4.5 years)) with prenatal DNAm and postpartum EPDS measures. The average EPDS score was 5.4, and 17.5% of women endorsed a positive lifetime history of MD. Neither MD history nor EPDS score was significantly associated with gestational age at delivery. The AYATS sample included N=150 participants (75 MZ European-American twin pairs, 72% female, mean age = 17.0 years (sd = 1.3 years)). Approximately 26% of the participants endorsed a lifetime history of MD (mean age of onset 14.7 years).

### 3.2 Predictive accuracy

Summary information for all models tested with the PREG study and AYATS can be found in Table 1. The predictive strength of the algorithm was modest (AUC: 0.62 [CI:0.45-0.79] in the full PREG sample; Figure 1a), and increasing the EPDS threshold to 15 did not improve the AUC; however, the algorithm exhibited high accuracy predicting high postpartum depressive symptom load in the subset of PREG participants with a lifetime history of MD (AUC:0.94 [CI:0.79-1.00]; Figure 1b). Sensitivity analyses in the full PREG sample and the African-American subset suggest that the accuracy of the algorithm for predicting high postpartum depressive symptom load within the first seven months postpartum may not be strongly affected by age. While the AUC point estimates for the older subset of the full PREG sample and the African-American subsets are larger than those for the younger subsets, the 95% confidence intervals (95% CI) overlap substantially (0.83 [95% CI: 0.59-.90] versus 0.74 [95% CI: 0.55-0.93] and 0.83 [95% CI: 0.63-1.00] and 0.78 [95% CI: 0.56-0.99] for the full sample and African-American subset, respectively).

**Figure 1:**
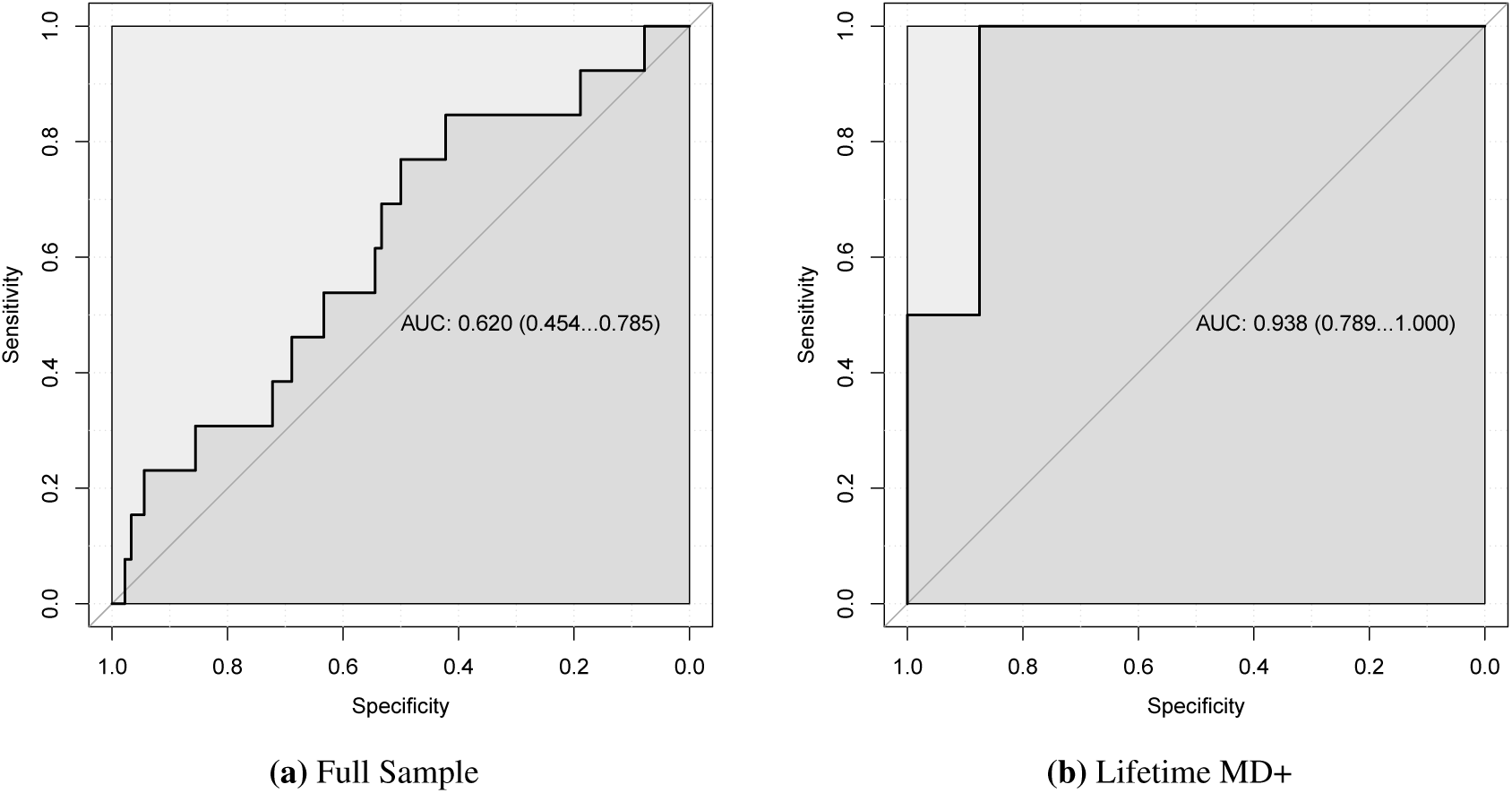
Area under the receiver operator characteristic curve predicting probable postpartum depression in PREG. The algorithm performed significantly better in the subset of PREG participants who endorsed having a positive lifetime history of major depression (MD) (**1b**) compared to the full PREG sample (**1a**).

**Figure 2:**
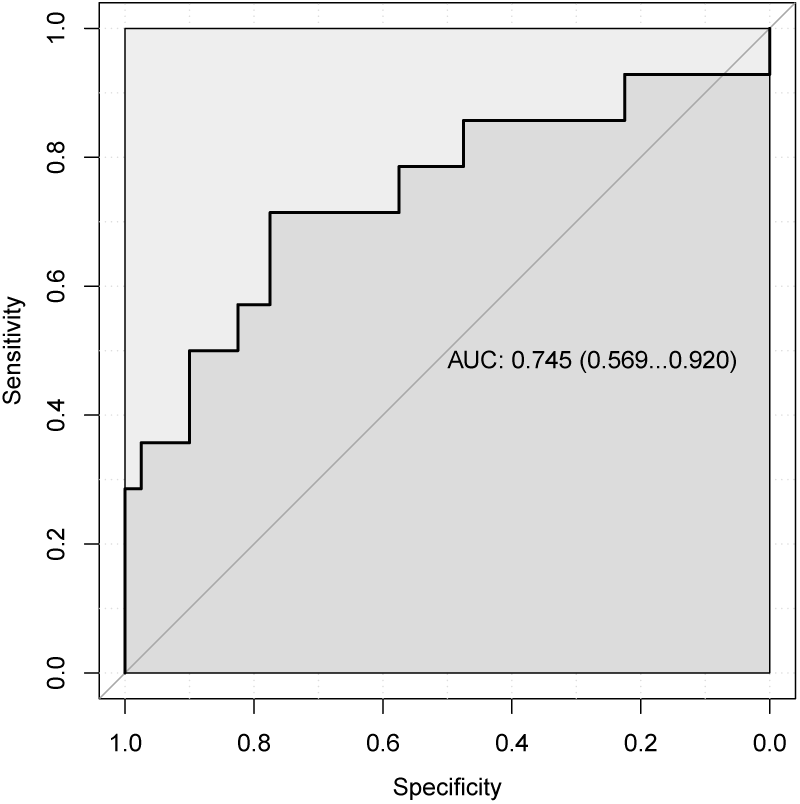
Area under the receiver operator characteristic curve predicting lifetime history of major depression in female participants from the AYATS sample.

The biomarker model demonstrated similar levels of accuracy for predicting lifetime history of MD in men using data from AYATS (0.83 [95% CI: 0.63-1.00]; average between twin 1 (T1) and twin 2 (T2) compared to the replication attempt using data from the Franconian Maternal Health Evaluation Studies (FRAMES; 0.81 [95% CI: 0.68-0.93]); ^32^ however, the AUC estimates were lower for predicting lifetime history of MD in adolescent females, and the 95% CIs for one subset of female twins included 0.50 (see Table 1).

## 4. Discussion

The purpose of these analyses were to assess the generalizability, predictive validity, and specificity of a DNAm-based model for prospectively predicting PPD from prenatal maternal blood. Collectively, the findings suggest the model identified individuals who have a positive lifetime history of MD and, for those individuals, can predict who will go on to develop postpartum depression. Further, the results suggest that the model may not be specific to pregnancy-related depression. Reasonably high AUC point estimates were obtained from a ROC analysis for both the male and female subsets of AYATS participants (0.85 and 0.74, respectively). Taken together, it is possible that the model correctly identifies women at risk for high levels of postpartum depression because those are the women with the greatest severity of lifetime MD. It also is possible that this model works especially well in individuals with a history of MD. In hindsight, a careful review of previous work developing and testing this model further supports the notion that the biomarker model may be primed to work best in individuals with a history of depressive psychopathology. First, the DNAm biomarkers were selected based on their joint predictive power in a clinical sample of participants with histories of either major depression or bipolar disorder (BP). ^11^ This clinical bias continued in the replication study, which used primarily cohorts enriched for depressive psychopathology (i.e., the Genetics of Recurrent Early Onset Depression (GenRED; ^33^ 50% MD) and the Prospective Gene Expression Postpartum Cohort from the Emory Women’s Mental Health Program ^34^ (60% MD, 40% BP; see Table 2). ^12^ Second, the analyses of DNAm remodeling trajectories in the Franconian Maternal Health Evaluation Studies (FRAMES) ^35^ and GenRED suggest that women with a history of PPD follow distinctly different DNAm patterns over time in both samples. This observation suggests that experiencing an episode of PPD may lead to subsequent epigenetic scarring in genomic regions associated with hormone responsiveness. It is possible that similar kinds of DNAm scars occur as a result of non-pregnancy-related MD episodes. Taken together, these results cumulatively support the hypothesis that MD episodes can evoke chronic perturbances in the DNA methylome and that interindividual variation in DNAm changes may explain why some women with a history of MD are at greater risk to develop PPD compared to other women with previous MD episodes.

It remains unclear if the DNAm values at these sites reflect risk for postpartum depression, a consequence of prior depressive episodes, a mixture of both, or something else entirely that correlates with risk for MD. More work is needed to understand the biological pathways and functional consequences associated with DNA methylation states at these two genomic loci. Carefully constructed longitudinal prospective studies will be necessary to disentangle the direction of causation between MD episodes that onset before, during, and after pregnancy and the DNA methylation patterns and remodeling associated with each. Addressing these questions along with concerns about generalizability across age and ethnicities is a critical step to applying this research to clinical practice. An accurate predictive algorithm for perinatal depression could have major implications for maternal-offspring health. Both perinatal depression and subclinical perinatal depressive symptoms have been associated with adverse maternal-infant health outcomes. ^4^ Recognizing maternal depression during pregnancy can be difficult due to the overlap between somatic signs of depression and typical pregnancy feelings/experiences (e.g., difficulty sleeping, changes in appetite), which is precisely why a biomarker test would be invaluable.

The strengths of this study include its prospective sampling of DNAm and multiple measures of depressive psychopathology. Moreover, the PREG study is demographically diverse in that it includes both African-American and European-American women from a wide range of ages (18-40) and pregnancy histories. Furthermore, data from a modest sample of never-pregnant adolescent participants was included to test if an episode of early-onset MD in the absence of any pregnancy was sufficient to be identified by the predictive algorithm. Additionally, the main analyses of this work were preregistered, which enables readers to distinguish planned tests from *posthoc* follow up analyses. However, the results should be considered in the context of study limitations. Most importantly, the replication analysis centers on using the general model described in Osborne (2016); however, without the the same coefficients that were used in that analysis, the exact model cannot be tested here. Secondly, the two samples used in these analyses (especially the twin sample) were of modest size, which contributed to large confidence intervals around AUC estimates. Also, both studies relied on a self-report version of the CIDI-SF to assess lifetime history of depressive psychopathology, and the PREG study used the EPDS to assess current levels of perinatal depressive symptoms. While self-report instruments have been associated with inflated estimates of MDP prevalence, there is evidence that EPDS is one of a few self-report instruments that has good sensitivity and specificity but does not overestimate the prevalence of perinatal depressive episodes meeting clinical thresholds compared to structured clinical interviews. ^36^ This detail is important because it suggests that using the EPDS may mitigate the issue of misclassification. Finally, antidepressant medication use was not assessed in the PREG cohort. That said, in both the original ^11^ and replication ^12^ studies, antidepressant use did not affect the AUC estimates of the algorithm’s predictive validity.

## Data Availability

The preregistration document and R code used to analyze the data and generate figures is available on the Open Science Framework (OSF) project landing page (https://osf.io/7dsf9). Data sharing from the Pregnancy, Race, Environment, Genes (PREG) study and Adolescent and Young Adult Twin Study are limited by Institutional Review Board (IRB) agreements and participant consent forms, which restrict openly sharing individual-level DNA methylation measures. Individuals interested in PREG or AYATS data access or collaboration are encouraged to contact Dr. Timothy P. York or Dr. Roxann Roberson-Nay, respectively.

## 5. Declarations

The authors have no conflicts of interest to declare.

## 5.1 Acknowledgements

The Pregnancy, Race, Environment, Genes (PREG) longitudinal study and its postpartum extension were supported by the NIHMD (P60MD002256, PI: York, Strauss), The John and Polly Sparks Foundation and Brain and Behavior Research Foundation (24712, PI: York), American Nurses Foundation Research Grant (5232, PI: Kinser), Virginia Commonwealth University Center for Clinical and Translational Research Endowment Fund (6-40595, PI: Kinser). The Adolescent and Young Adult Twin Study was supported by a NARSAD Independent Investigator Award from the Brain and Behavior Research Foundation and by the NIMH (R01MH101518) to RRN. DL is supported by NIMH T32MH020030 (PI: M. Neale). Both studies used REDCap to administer self-report questionnaires, and the use of REDCap was supported by Clinical and Translational Science Award (CTSA) award No. UL1TR000058 from the National Center for Advancing Translational Sciences. Its contents are solely the responsibility of the authors and do not necessarily represent official views of the National Center for Advancing Translational Sciences or the National Institutes of Health.

## 5.2 Informed consent and study approval

The Virginia Commonwealth University Institutional Review Board (VCU IRB) approval and written participant consent was established for both the PREG study (14000) and AYATS (HM15348). Written parental consent was obtained for AYATS participants who were under the age of 18 years old.

## 5.4 Author contributions

DL planned and carried out the analysis and wrote the initial manuscript draft. TPY and PK conceived of and secured funding for the Pregnancy, Race, Environment, Genes (PREG) study and the postpartum extension study, respectively. RRN conceived of the Adolescent and Young Adult Twin Study (AYATS). All co-authors reviewed the manuscript and approved the final version.

## 5.5 Abbreviations

AUC: Area under the curve
AYATS: Adolescent and Young Adult Twin Study
DNAm: DNA methylation
EPDS: Edinburgh Postnatal Depression Scale
MD: Major depression
MDP: Major depression in the peripartum
MZ: Monozygotic
PPD: Postpartum depression
PREG: Pregnancy, Race, Environment, Genes Study
ROC: Receiver operator characteristic

